# Does Metformin Decrease Mortality in Patients with Type 2 Diabetes Mellitus Hospitalized for COVID-19? A Multivariable and Propensity Score-adjusted Meta-analysis

**DOI:** 10.1101/2022.04.03.22273353

**Authors:** Zhiyuan Ma, Mahesh Krishnamurthy

## Abstract

**Aims:** Coronavirus disease 2019 (COVID-19) is a new pandemic that the entire world is facing since December of 2019. Increasing evidence has shown that metformin is linked to favorable outcomes in patients with COVID-19. The aim of this study was to address whether outpatient or inpatient metformin therapy offers low in-hospital mortality in patients with type 2 diabetes mellitus hospitalized for COVID-19.

**Methods:** We searched studies published in PubMed, Embase, Google Scholar and Cochrane Library up to October 1, 2021. Raw event data extracted from individual study were pooled using the Mantel-Haenszel approach. Odds ratio (OR) or hazard ratio (HR) adjusted for covariates that potentially confound the association using multivariable regression or propensity score matching was pooled by the inverse-variance method. Random effect models were applied for meta-analysis due to variation among studies.

**Results:** Nineteen retrospective observational studies were selected. The pooled unadjusted OR for outpatient metformin therapy and in-hospital mortality was 0.54 (95% CI, 0.42-0.68), whereas the pooled OR adjusted with multivariable regression or propensity score matching was 0.72 (95% CI, 0.47-1.12). The pooled unadjusted OR for inpatient metformin therapy and in-hospital mortality was 0.19 (95% CI, 0.10-0.36), whereas the pooled adjusted HR was 1.10 (95% CI, 0.38-3.15).

**Conclusions:** Our results suggest that there is a significant reduction of in-hospital mortality with metformin therapy in patients with type 2 diabetes mellitus hospitalized for COVID-19 in the unadjusted analysis, but this mortality benefit does not retain after adjustments for confounding bias.

## Introduction

Coronavirus disease 2019 (COVID-19),caused by the novel severe acute respiratory syndrome coronavirus 2 (SARS-CoV-2), is a new pandemic, leading to high mortality [1–5]. One of risk factors linked to worse outcomes for COVID-19 is preexisting type 2 diabetes mellitus [6]. Patients with diabetes hospitalized with COVID-19 are two- to three-fold more likely to be admitted into intensive care units than that of non-diabetics and the mortality rate is at least doubled [7–10].

There seems two distinct but overlapping pathologic phases in response to SARS-CoV-2 infection: the initial viral response phase triggered by the virus itself and the subsequent inflammatory phase triggered by the host response [11]. Accumulating evidence has shown cytokine storm syndrome in patients with severe COVID-19. Hence, agents that alleviate hyper-inflammation in COVID-19 patients could be beneficial. Metformin has been shown to modulate the immune responses and restore immune homeostasis in immune cells via AMPK-dependent mechanisms [12]. Historically, metformin was used during the treatment of influenza outbreak, due to its host-directed anti-viral properties [13]. In light of the pathogenesis of SARS-CoV-2, several possible beneficial effects of metformin therapy in COVID-19 patients with pre-existing type 2 diabetes mellitus have been speculated, such as anti-inflammatory effects [14], reduction in neutrophils [15], increasing the cellular pH to inhibit viral infection, interfering with the endocytic cycle and reversing established lung fibrosis [16]. Conversely, metformin therapy has also been challenged for the potential risk for lactic acidosis, particularly in the cases of multi-organ failure and for promoting SARS-CoV-2 infection by possibly increasing in ACE2 availability in the respiratory tract [17].

Accumulating evidence from retrospective studies suggests that treating COVID-19 patients with pre-existing type 2 diabetes mellitus with metformin may not be harmful, but even endows a protective effect and offers a low mortality. Due to the inherent nature of retrospective studies, biases are likely to confound the association between metformin therapy and outcomes. In this study, to limit the confounding bias, we aimed to address whether outpatient or inpatient metformin therapy offers low in-hospital mortality in patients with type 2 diabetes mellitus hospitalized for COVID-19 by multivariable and propensity score-adjusted meta-analysis.

## Methods

### Strategy of literature search for meta-analysis

A systemic literature search of studies published in English was performed in PubMed, Embase, Google Scholar and Cochrane Library up to October 1, 2021 using the key words ‘metformin or biguanides’, ‘SARS-CoV2 or COVID-19’ and ‘mortality’. All abstracts were screened and the ‘related articles’ function was employed to broaden the search on appropriate abstracts including meta-analysis. Full texts were then retrieved and reviewed. The meta-analysis was in adherence to the principles outlined by the Preferred Reporting Items for Systematic Reviews and Meta-Analyses (PRISMA). The study was registered on the PROSPERO (CRD42021284113).

### Study selection and risk of bias assessment for meta-analysis

Two reviewers independently extracted the following data from each study: first author, year of publication, study population characteristics (age and gender), study size, study design, country, definition of mortality, number of metformin users, effect size including odds ratios (ORs) or hazard ratios (HRs), confounding adjustment, and timing of metformin use. Studies included in the analysis met: 1) comparing the effect of metformin use prior to admission or during hospitalization on COVID-19 patients with type 2 diabetes; 2) the raw data on events can be extracted from the manuscript or supplementary materials or effect sizes (OR, HR) were reported; and 3) outcomes include in-hospital mortality. Studies were discarded, if: 1) duplicated studies from the same Registry or dataset; 2) studies without a control group or without in-hospital mortality reported. 3) total patients with type 2 diabetes in the study < 50; 4) studies that were not published in English. Discrepancies were resolved by discussion. The Risk of Bias in the included studies was assessed using the Newcastle-Ottawa Scale [18].

### Data syntheses and statistical analysis

To assess whether metformin use offers low in-hospital mortality by comparing and integrating the results of different studies, meta-analysis was carried out as described previously [19]. OR was used as the main summary statistics. The OR represents the odds of a death event occurring in the metformin group in comparison to the non-metformin group. The point of estimate of the OR is considered statistically significant at the P < 0.05 level if the 95% confidence interval does not include the value 1. Raw event data extracted from individual study were pooled using the Mantel-Haenszel approach and effect size was pooled using the inverse-variance method. In our study, random effect models were employed due to variation between studies. Heterogeneity between studies was investigated by the standard chi-squared Q-test. Publication bias was assessed by Funnel plots and *Peters* test. In addition, a sensitivity analysis was performed by excluding one study at a time to assess the robustness of the results. All analyses were conducted with R (version 3.6.1) and R package ‘meta’.

## Results

### Study selection and characteristics

In the literature search, 122 studies were identified. After studies were carefully reviewed based on the inclusion and exclusion criteria, 19 retrospective observational studies were selected for further meta-analysis of metformin use and in-hospital mortality. The flowchart for the study selection is shown in Figure 1. Of these 19 studies, 14 studies reported on the association of outpatient metformin use and in-hospital mortality [20–33], 4 studies investigated the link between inpatient metformin use and in-hospital mortality [34–37], and 1 study probed the association of both outpatient and inpatient metformin use and in-hospital mortality [38]. There were 13 studies with extractable raw event data. According to adjustment for confounding bias, 14 studies performed multivariable regression or propensity scoring matching, in which a propensity score was calculated based on logistic regression to control for confounding factors that could influence the metformin use. The characteristics of the included studies are summarized in Table 1.

**Table 1.**
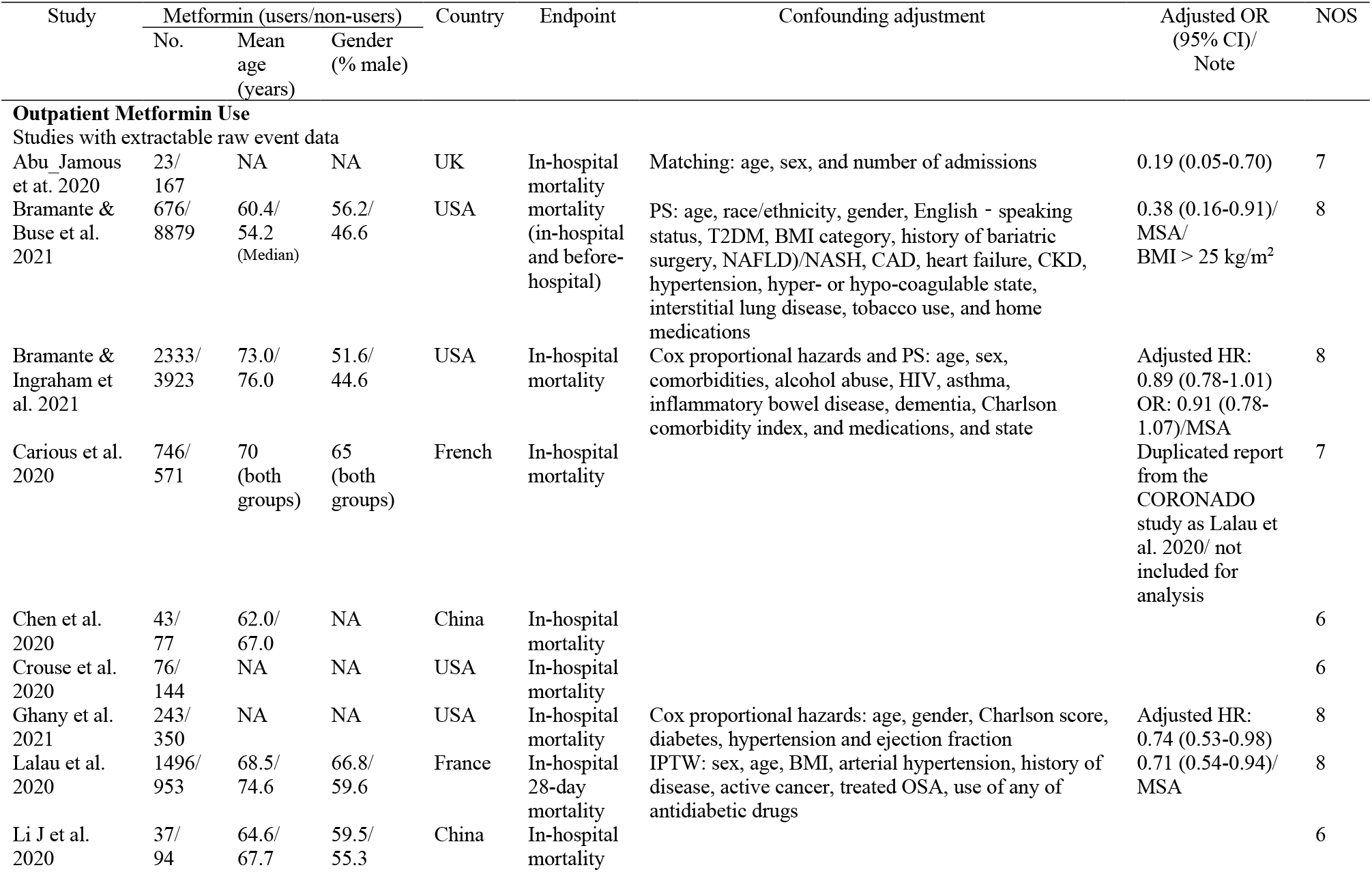
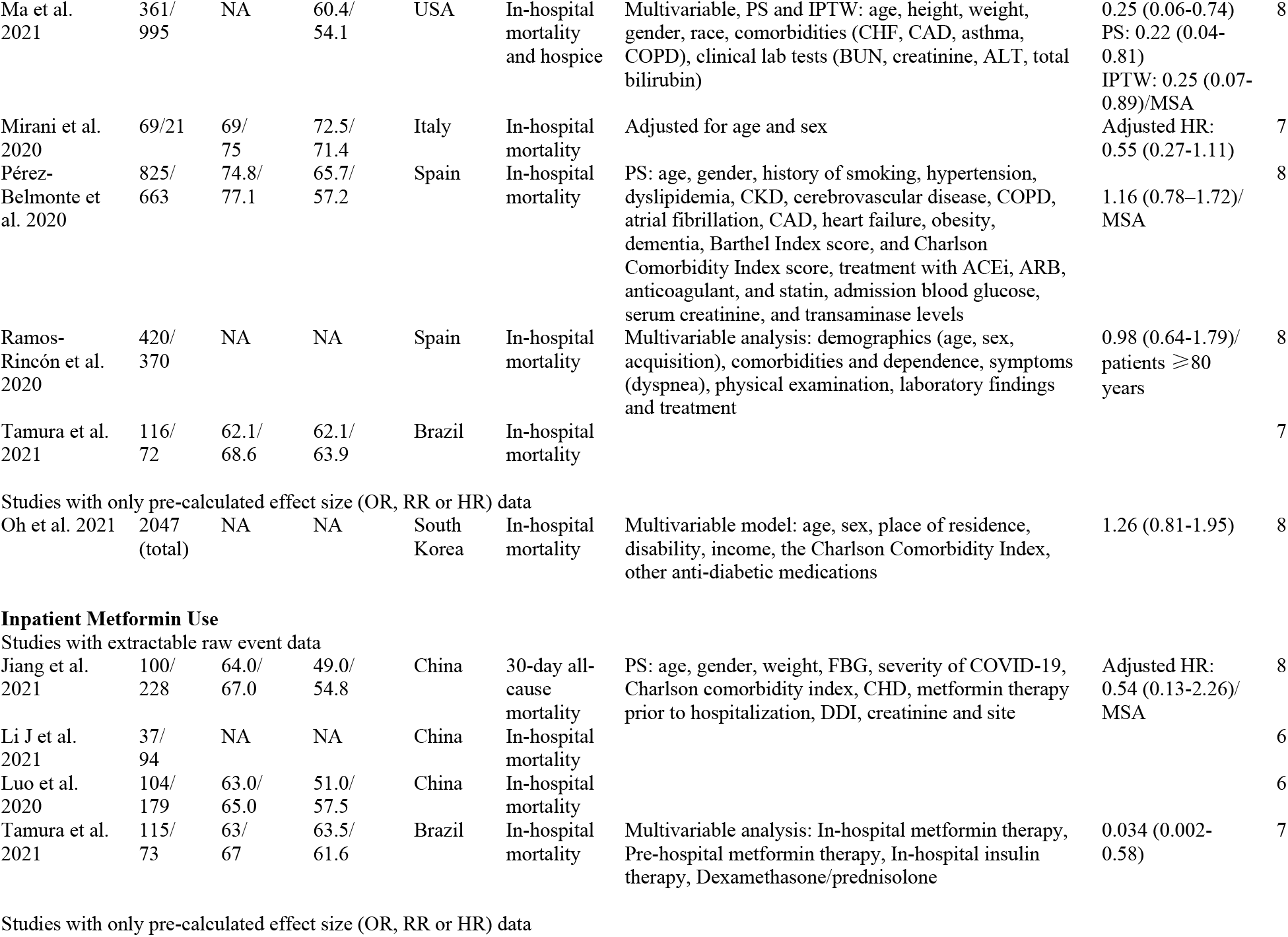
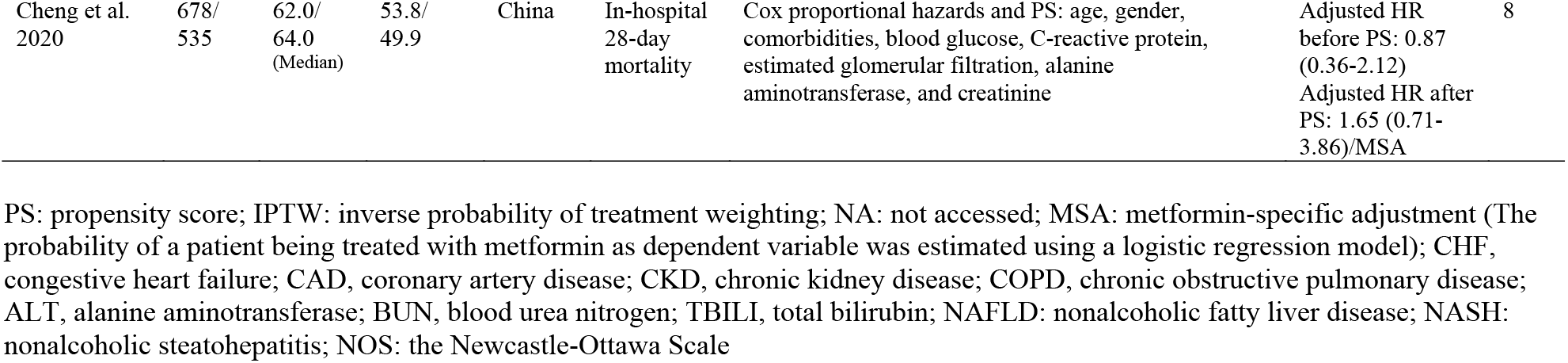
Characteristics of studies comparing mortality in metformin users vs non-users in patients with type 2 diabetes hospitalized for COVID-19.

| Study | Metformin (users/non-users) |  |  | Country | Endpoint | Confounding adjustment | Adjusted OR<br>(95% CI)/<br>Note | NOS |
| --- | --- | --- | --- | --- | --- | --- | --- | --- |
|  | No. | Mean<br>age<br>(years) | Gender<br>(% male) |  |  |  |  |  |
| Outpatient Metformin Use |  |  |  |  |  |  |  |  |
| Studies with extractable raw event data |  |  |  |  |  |  |  |  |
| Abu_Jamous<br>et at. 2020 | 23/<br>167 | NA | NA | UK | In-hospital<br>mortality | Matching: age, sex, and number of admissions | 0.19 (0.05-0.70) | 7 |
| Bramante &<br>Buse et al.<br>2021 | 676/<br>8879 | 60.4/<br>54.2<br>(Median) | 56.2/<br>46.6 | USA | mortality<br>(in-hospital<br>and before-<br>hospital) | PS: age, race/ethnicity, gender, English - speaking<br>status, T2DM, BMI category, history of bariatric<br>surgery, NAFLD)/NASH, CAD, heart failure, CKD,<br>hypertension, hyper- or hypo-coagulable state,<br>interstitial lung disease, tobacco use, and home<br>medications | 0.38 (0.16-0.91)/<br>MSA/<br>BMI > 25 kg/m² | 8 |
| Bramante &<br>Ingraham et<br>al. 2021 | 2333/<br>3923 | 73.0/<br>76.0 | 51.6/<br>44.6 | USA | In-hospital<br>mortality | Cox proportional hazards and PS: age, sex,<br>comorbidities, alcohol abuse, HIV, asthma,<br>inflammatory bowel disease, dementia, Charlson<br>comorbidity index, and medications, and state | Adjusted HR:<br>0.89 (0.78-1.01)<br>OR: 0.91 (0.78-1.07)/MSA | 8 |
| Carious et al.<br>2020 | 746/<br>571 | 70<br>(both<br>groups) | 65<br>(both<br>groups) | French | In-hospital<br>mortality |  | Duplicated report<br>from the<br>CORONADO<br>study as Lalau et<br>al. 2020/ not<br>included for<br>analysis | 7 |
| Chen et al.<br>2020 | 43/<br>77 | 62.0/<br>67.0 | NA | China | In-hospital<br>mortality |  |  | 6 |
| Crouse et al.<br>2020 | 76/<br>144 | NA | NA | USA | In-hospital<br>mortality |  |  | 6 |
| Ghany et al.<br>2021 | 243/<br>350 | NA | NA | USA | In-hospital<br>mortality | Cox proportional hazards: age, gender, Charlson score,<br>diabetes, hypertension and ejection fraction | Adjusted HR:<br>0.74 (0.53-0.98) | 8 |
| Lalau et al.<br>2020 | 1496/<br>953 | 68.5/<br>74.6 | 66.8/<br>59.6 | France | In-hospital<br>28-day<br>mortality | IPTW: sex, age, BMI, arterial hypertension, history of<br>disease, active cancer, treated OSA, use of any of<br>antidiabetic drugs | 0.71 (0.54-0.94)/<br>MSA | 8 |
| Li J et al.<br>2020 | 37/<br>94 | 64.6/<br>67.7 | 59.5/<br>55.3 | China | In-hospital<br>mortality |  |  | 6 |
| Ma et al.<br>2021 | 361/<br>995 | NA | 60.4/<br>54.1 | USA | In-hospital<br>mortality<br>and hospice | Multivariable, PS and IPTW: age, height, weight,<br>gender, race, comorbidities (CHF, CAD, asthma,<br>COPD), clinical lab tests (BUN, creatinine, ALT, total<br>bilirubin) | 0.25 (0.06-0.74)<br>PS: 0.22 (0.04-0.81)<br>IPTW: 0.25 (0.07-0.89)/MSA | 8 |
| Mirani et al.<br>2020 | 69/21 | 69/<br>75 | 72.5/<br>71.4 | Italy | In-hospital<br>mortality | Adjusted for age and sex | Adjusted HR:<br>0.55 (0.27-1.11) | 7 |
| Pérez-Belmonte et<br>al. 2020 | 825/<br>663 | 74.8/<br>77.1 | 65.7/<br>57.2 | Spain | In-hospital<br>mortality | PS: age, gender, history of smoking, hypertension,<br>dyslipidemia, CKD, cerebrovascular disease, COPD,<br>atrial fibrillation, CAD, heart failure, obesity,<br>dementia, Barthel Index score, and Charlson<br>Comorbidity Index score, treatment with ACEi, ARB,<br>anticoagulant, and statin, admission blood glucose,<br>serum creatinine, and transaminase levels | 1.16 (0.78–1.72)/<br>MSA | 8 |
| Ramos-Rincón et al.<br>2020 | 420/<br>370 | NA | NA | Spain | In-hospital<br>mortality | Multivariable analysis: demographics (age, sex,<br>acquisition), comorbidities and dependence, symptoms<br>(dyspnea), physical examination, laboratory findings<br>and treatment | 0.98 (0.64-1.79)/<br>patients ≥80<br>years | 8 |
| Tamura et al.<br>2021 | 116/<br>72 | 62.1/<br>68.6 | 62.1/<br>63.9 | Brazil | In-hospital<br>mortality |  |  | 7 |
| Studies with only pre-calculated effect size (OR, RR or HR) data |  |  |  |  |  |  |  |  |
| Oh et al. 2021 | 2047<br>(total) | NA | NA | South<br>Korea | In-hospital<br>mortality | Multivariable model: age, sex, place of residence,<br>disability, income, the Charlson Comorbidity Index,<br>other anti-diabetic medications | 1.26 (0.81-1.95) | 8 |
| **Inpatient Metformin Use** |  |  |  |  |  |  |  |  |
| Studies with extractable raw event data |  |  |  |  |  |  |  |  |
| Jiang et al.<br>2021 | 100/<br>228 | 64.0/<br>67.0 | 49.0/<br>54.8 | China | 30-day all-<br>cause<br>mortality | PS: age, gender, weight, FBG, severity of COVID-19,<br>Charlson comorbidity index, CHD, metformin therapy<br>prior to hospitalization, DDI, creatinine and site | Adjusted HR:<br>0.54 (0.13-2.26)/<br>MSA | 8 |
| Li J et al.<br>2021 | 37/<br>94 | NA | NA | China | In-hospital<br>mortality |  |  | 6 |
| Luo et al.<br>2020 | 104/<br>179 | 63.0/<br>65.0 | 51.0/<br>57.5 | China | In-hospital<br>mortality |  |  | 6 |
| Tamura et al.<br>2021 | 115/<br>73 | 63/<br>67 | 63.5/<br>61.6 | Brazil | In-hospital<br>mortality | Multivariable analysis: In-hospital metformin therapy,<br>Pre-hospital metformin therapy, In-hospital insulin<br>therapy, Dexamethasone/prednisolone | 0.034 (0.002-0.58) | 7 |
| Studies with only pre-calculated effect size (OR, RR or HR) data |  |  |  |  |  |  |  |  |
| Cheng et al.<br>2020 | 678/<br>535 | 62.0/<br>64.0<br>(Median) | 53.8/<br>49.9 | China | In-hospital<br>28-day<br>mortality | Cox proportional hazards and PS: age, gender,<br>comorbidities, blood glucose, C-reactive protein,<br>estimated glomerular filtration, alanine<br>aminotransferase, and creatinine | Adjusted HR<br>before PS: 0.87<br>(0.36-2.12)<br>Adjusted HR after<br>PS: 1.65 (0.71-3.86)/MSA | 8 |

PS: propensity score; IPTW: inverse probability of treatment weighting; NA: not accessed; MSA: metformin-specific adjustment (The probability of a patient being treated with metformin as dependent variable was estimated using a logistic regression model); CHF, congestive heart failure; CAD, coronary artery disease; CKD, chronic kidney disease; COPD, chronic obstructive pulmonary disease; ALT, alanine aminotransferase; BUN, blood urea nitrogen; TBILI, total bilirubin; NAFLD: nonalcoholic fatty liver disease; NASH: nonalcoholic steatohepatitis; NOS: the Newcastle-Ottawa Scale

**Figure 1.**
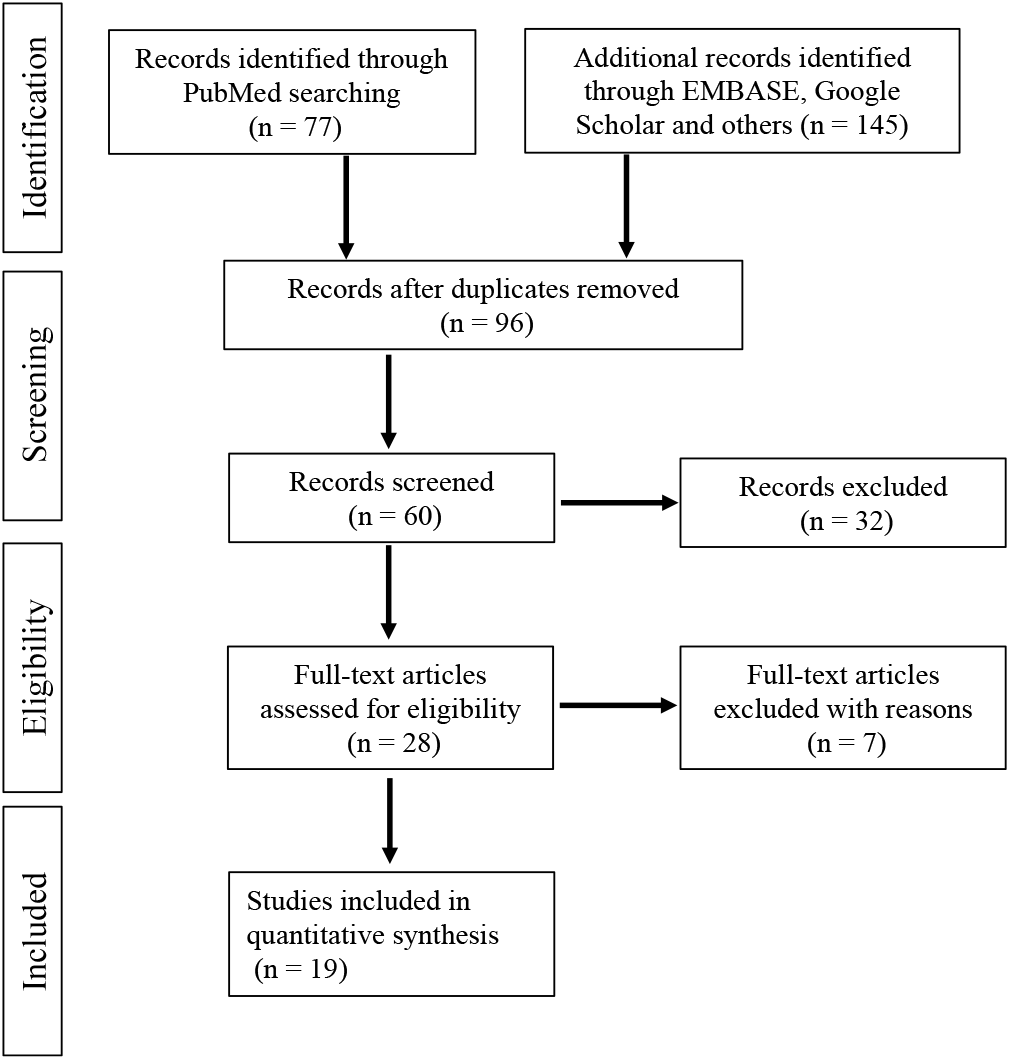
Flowchart illustration of the study selection.

### Outpatient metformin use and in-hospital mortality

There were 15 studies with or without attempt to adjust for confounding examining the association between outpatient metformin use and in-hospital mortality in patients with type 2 diabetes hospitalized for COVID-19. Due to the inherent nature of retrospective observational studies, to better probe the in-hospital mortality benefits and risks for outpatient metformin use, we set out to perform the meta-analysis using two approaches.

First, meta-analysis of 13 studies with extractable raw event data from 6,718 metformin and 16,708 non-metformin users revealed that outpatient metformin use was associated with a significant reduction of in-hospital death (Figure 2A) with a pooled unadjusted OR of 0.54 (95% CI, 0.42-0.68; *I*^*2*^ = 73%). Publication bias was assessed using a funnel plot (Figure 2B) which showed the 13 studies used in our meta-analysis. The scatter plot resembles an inverted funnel (the 95% confidence interval), in which the treatment effects estimated from individual studies on the horizontal axis (OR), against a measure of study size on the vertical funnel (SE [log OR]). *Peters* test was performed to quantitively assess publication bias and revealed publication bias (p = 0.029)

**Figure 2.**
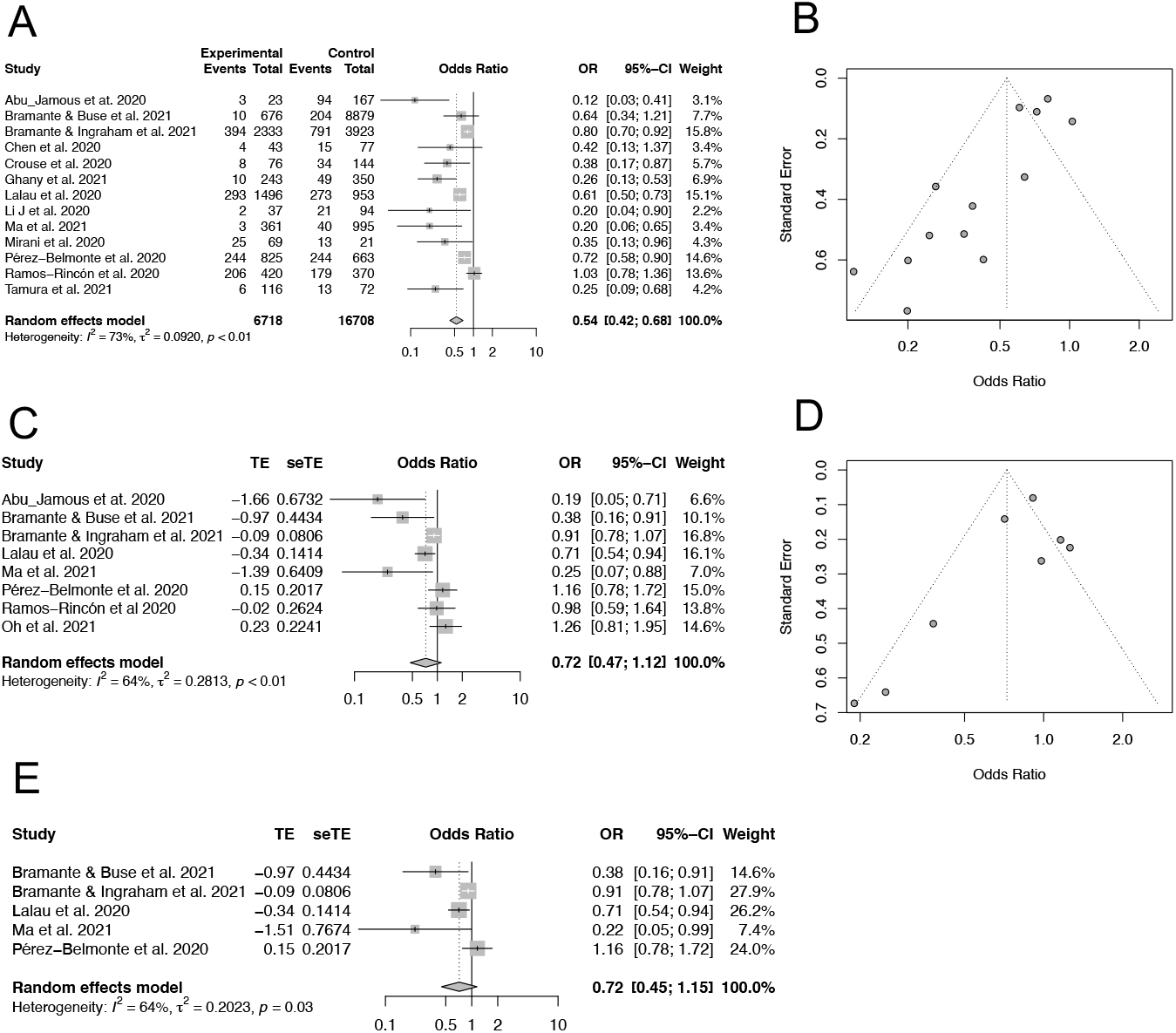
The association of outpatient metformin use and in-hospital mortality in patients with COVID-19 and pre-existing T2DM. (A) Meta-analysis of 13 selected studies with extractable raw event data using the Mantel-Haenszel approach. (B) Funnel plot test of 13 studies included in panel A. (C) Meta-analysis of 8 selected studies with multivariable or propensity score-adjusted OR using the inverse-variance method to pool effect sizes. (D) Funnel plot of 8 studies included in panel C. (E) Meta-analysis of OR for in-hospital mortality in 5 studies with propensity score matched for metformin use by the inverse-variance method.

Second, in contrast, meta-analysis of 8 studies with adjusted OR using either multivariable regression or propensity score matching demonstrated a pooled OR of 0.72 (95% CI, 0.47-1.12; *I*^*2*^ = 64%), suggesting that there was a nonsignificant trend toward a decrease in-hospital mortality associated with outpatient metformin use (Figure 2C). The funnel plot for the included 8 studies showed an approximate symmetric distribution (Figure 2D). *Peters* test was not performed due to small number of studies (n < 10).

To better limit confounding bias due to baseline characteristics, several studies conducted analyses with propensity score matching approaches, in which propensity score was derived as the probability of being treated with metformin based on the participant’s observed covariates between metformin and non-metformin users. Likewise, subgroup analysis of propensity scored-matched studies, outpatient metformin use was not associated with a statistically significant reduction of in-hospital mortality with a pooled OR of 0.72 (95% CI, 0.45-1.15; *I*^*2*^ = 64%) (Figure 2E). In addition, to determine the impact of individual studies on pooled effects, sensitivity analysis was performed by excluding one study a time. Neither the pooled ORs in studies with extractable raw event data (Supplementary figure 1A) showed significant differences, nor was in studies with adjusted OR (Supplementary figure 1B). These results indicate that outpatient metformin use is associated with a significant reduction of in-hospital death in unadjusted raw data, but it was not statistically significant after adjustments for confounding.

### Inpatient metformin use and in-hospital mortality

There were a few studies mainly from China investigating the association between inpatient metformin use and in-hospital mortality in patients with type 2 diabetes hospitalized for COVID-19. We performed the similar analyses with extractable raw data and adjusted OR. Meta-analysis of 4 studies with extractable raw event data from 356 metformin and 574 non-metformin users showed that inpatient metformin use was associated with a significant reduction of in-hospital death (Figure 3A) with a pooled unadjusted OR of 0.19 (95% CI, 0.10-0.36; *I*^*2*^ = 0%). However, meta-analysis of 2 studies with adjusted HR demonstrated a pooled HR of 1.10 (95% CI, 0.38-3.15; *I*^*2*^ = 43%), suggesting there is no statistically significant association between inpatient metformin use and in-hospital mortality (Figure 3B). No sensitivity or subgroup studies were performed due to small number of studies available.

**Figure 3.**
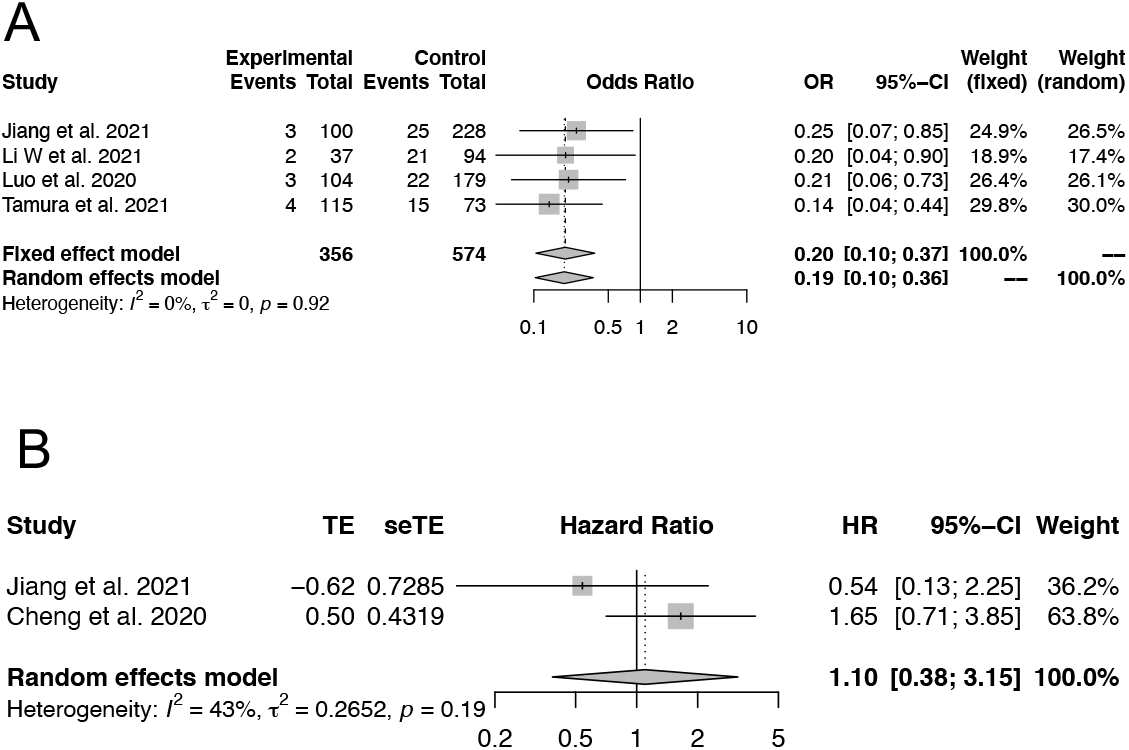
The association of inpatient metformin use and in-hospital mortality in patients with COVID-19 and pre-existing T2DM. (A) Meta-analysis of 4 selected studies with extractable raw event data using the Mantel-Haenszel approach. (B) Meta-analysis of 2 selected studies with multivariable and propensity score-adjusted HR using the inverse-variance method to pool effect sizes.

## Discussion

Diabetes has been linked to poor outcomes in patients with COVID-19, so treatments that are effective, safe, and available immediately are urgently needed. Metformin is a the first-line therapeutic agent that is safe, effective, and inexpensive for type 2 diabetes mellitus. In the light of host-directed anti-viral properties and anti-inflammatory effects of metformin, many retrospective studies including our previous study [29] has explored whether metformin is beneficial in patients with COVID-19 and preexisting type 2 diabetes mellitus. To further compare and integrate the results of different studies and to increase statistical power, we set out to perform a meta-analysis. Prior to this meta-analysis, several meta-analyses [39–41] of the association between metformin therapy and severity and mortality were published, but they have issues. First, metformin therapy was not differentiated between outpatient and inpatient use, as oral hypoglycemic medications including metformin are usually held during hospitalization. Second, while study endpoints such as pre-defined mortality were somewhat different among studies, Effect sizes (OR or HR) of different mortality outcomes were combined in previous meta-analysis studies. Third, due to nature of retrospective studies, various effect sizes with adjustments for baseline characteristics were reported in several studies. However, unadjusted and adjusted effect sizes were mixed in different meta-analyses. Fourth, using pre-calculated standard error instead of raw event data may not be a good estimator of the precision of the binary effect size (OR or HR). Nevertheless, only pre-calculated OR was used in the previous meta-analyses.

In this study, we specifically addressed these issues by investigating the association between outpatient or inpatient metformin therapy and in-hospital mortality in patients with diabetes hospitalized for COVID-19. We also used extractable raw data instead of pre-calculated ORs or HRs to determine the pooled OR for the unadjusted analysis. We found that there was a significant reduction of in-hospital mortality with outpatient or inpatient metformin therapy in patients with type 2 diabetes mellitus hospitalized for COVID-19 in the unadjusted analysis, which is in line with previous meta-analyses [39–41]. Furthermore, evidence obtained from observational studies is an of great importance source for clinical practice where randomized clinical trials are unavailable or infeasible [42]. Unlike clinical trials through randomization to ensure patient characteristics are comparable across treatment and control groups, observational studies usually attempt to adjust for confounding by multivariable regression and propensity score matching. We took the advantage of these approaches and applied adjusted ORs or HRs into our meta-analysis. In contrast to the unadjusted analysis, neither outpatient nor inpatient metformin therapy was associated with decreased in-hospital mortality. When only the matched studies [21, 22, 27, 29, 32] were considered for outpatient metformin use and in-hospital mortality, a similar result was reached that there was no mortality benefit for outpatient metformin therapy in patient with diabetes hospitalized for COVD-19. These findings are in discordant with previous meta-analyses. Our results suggest that the mortality benefit observed in our unadjusted meta-analysis and previous reported meta-analyses could be due to confounding bias. Therefore, further randomized clinical trials are needed to investigate the clinical benefits of metformin therapy in patient with diabetes hospitalized for COVD-19.

Several limitations are present in our study. First, owing to lack of randomized clinical trials, all studies included for analysis were retrospective studies. Although efforts were made to balance and control for potential confounding factors by multiple variable adjustments and propensity score matching, due to the inherent nature of retrospective observational studies, residual confounders are likely to exist and could not be balanced. Therefore, even meta-analysis of studies with adjusted ORs o HRs could be misleading. Second, the size of the retrospective studies that were included in the meta-analysis varied considerably, resulting in moderate-to-high heterogeneity. Third, there were few studies investigating inpatient metformin use and in-hospital mortality. More studies are needed to perform a robust meta-analysis for inpatient metformin therapy and in-hospital mortality in patient with diabetes hospitalized for COVD-19.

In conclusion, our findings demonstrate that there is a significant reduction of in-hospital mortality with outpatient or inpatient metformin therapy in patients with type 2 diabetes mellitus hospitalized for COVID-19 in the unadjusted analysis, but this mortality benefit does not retain after adjustments for confounding bias. Therefore, further randomized clinical trials are needed to provide clinical evidence regarding metformin therapy and in-hospital mortality in patient with diabetes hospitalized for COVD-19.

## Data Availability

All data produced in the present work are contained in the manuscript

## Author Contributions

ZM and MK designed the study and wrote the manuscript. ZM and MK collected, performed, and analyzed data. All authors reviewed the results and approved the final version of the manuscript.

---

**Supplementary Figure 1.**
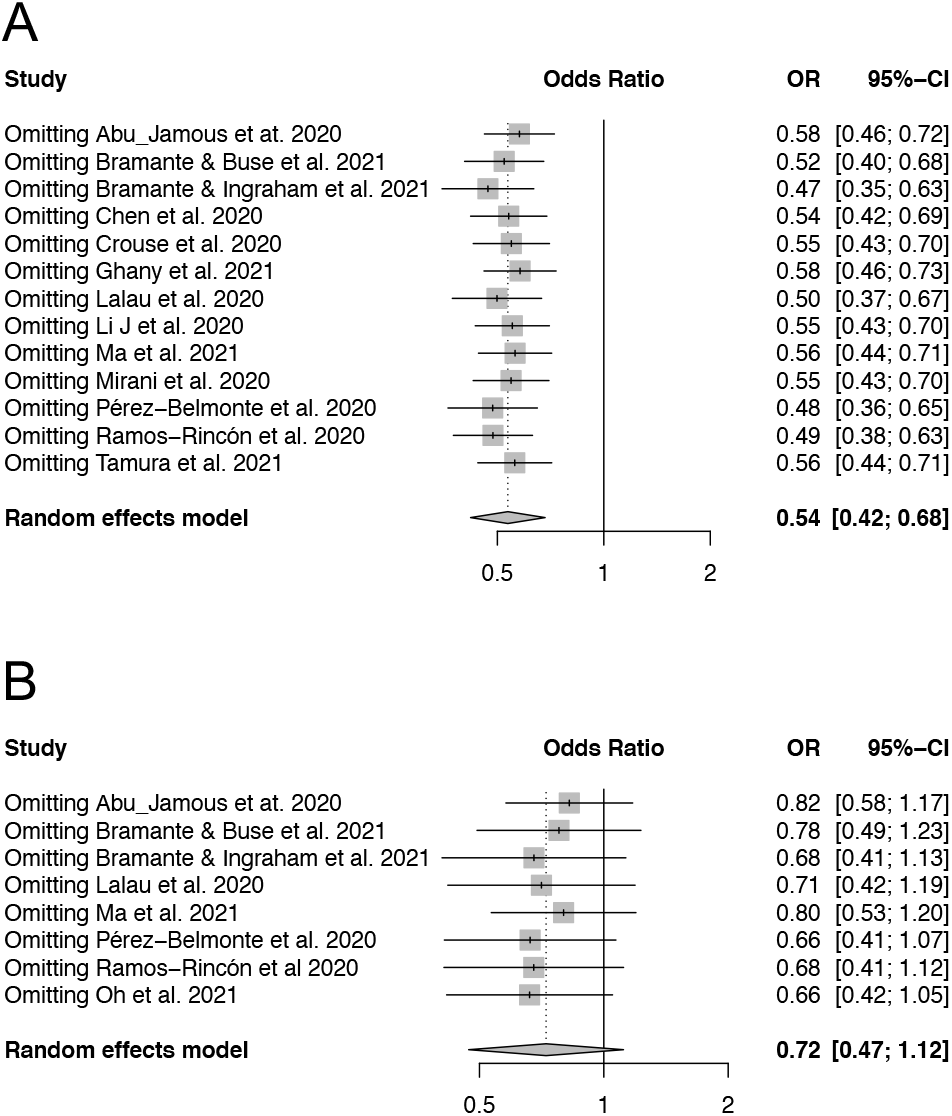
Sensitivity analysis of the studies included in meta-analysis in Figure 2A and 2C by excluding each study at a time.

